# Performance of the Reported and Intended Behavior Scale among Colombian Adolescents

**DOI:** 10.1101/2021.11.26.21266915

**Authors:** A. Campo-Arias, G. A. Ceballos-Ospino, E. Herazo

## Abstract

**Objective:** To establish the Reported and Intended Behavior Scale (RIBS) psychometric performance, a mental disorder-related stigma measurement, among Colombian adolescents.

**Methods:** A validation study was carried out with 350 students aged between 10 and 17, 53.7% of whom were girls. The RIBS has two sub-scales -reported behaviors and intended behaviors, with four items each. Frequencies were estimated for reported behaviors, whereas internal consistency (Cronbach’s alpha and McDonald’s omega) and confirmatory factor analysis (CFA) were measured for intended behaviors.

**Results:** The reported behavior sub-scale ranged from 10.0 to 24.9%, whereas the intended behavior sub-scale presented a Cronbach’s alpha of 0.88 (CI95% 0.86-0.90) and a McDonald omega of 0.88. For the CFA, KMO was 0.81; Bartlett chi squared, 771.1 (*df*=6, *p*=0.01); and Eigen value, 2.95 that explained 73.9% of the total variance. For the goodness-of-fit tests, chi squared was 21.9 (*df*=2, *p*=.001); RMSEA, 0.17 (CI90% 0.11-0.24); CFI, 0.97; TLI, 0.92; and SMSR, 0.03.

**Conclusions:** The RIBS can measure reported behaviors, and the intended behavior sub-scale shows high internal consistency. However, the dimensionality of the intended behavior sub-scale presents modest goodness-of-fit indexes. These findings need further replications.

## INTRODUCTION

The stigma-discrimination complex was recently introduced to integrate four highly interrelated concepts: stigma, stereotype, prejudice, and discrimination^1^. Stigma occurs when an individual or collective attribute, characteristic, condition, trait, or situation is given an unfavorable assessment (1, 2). Stereotypes are preconceived ideas of an attribute; this idea can be positive or negative and implies a simplification of the valuation of a person or group (1).

Prejudice occurs when the stereotype takes on a derogatory or pejorative connotation and is a quick judgment of or attitude towards the person as a whole (2). This judgment tends to consider the indicated trait only, omitting other aspects and the distinctive singularity of each person or any other information that could distort this almost automatic idea (2, 3). Stereotypes are also resistant to change, despite the availability of information that repeatedly denies this (4).

Finally, discrimination is configured when society validates prejudice and grants the person or groups status as second-class citizens, and is dismissive of a set of rights about those carrying the stigmatized characteristic (5). Stigma and discrimination happen simultaneously, which is why it is known as: “stigma-discrimination complex” (1, 6).

For several reasons, most people who meet the criteria for a mental disorder have suffered from a stigma-discrimination complex (1, 7). The mental disorder-related stigma-discrimination complex is ubiquitous among school children and young adolescents (8) and presents many adverse outcomes; it is linked to low self-esteem, few help-seeking behaviors, and even suicide among psychiatric patients (9).

Thus, the importance of the mental disorder-related stigma-discrimination complex has encouraged the design of instruments for its measurements, such as the Reported and Intended Behavior Scale (RIBS). The RIBS asks participants about reported and intended behavior in four different contexts: living with, working with, living near, and continuing a friendship with a person who meets the criteria for a mental disorder. The reported behavior sub-scale inquires about the frequency of experiences, and the intended behavior sub-scale measures attitudes as a construct. Evans-Lacko et al., in a sample of 403 adults, aged between 25 and 45, reported behaviors ranging from 24.8 to 43.9%, whereas the intended behavior sub-scale showed high internal consistency with a Cronbach’s alpha of 0.85 (10).

On the other hand, in English adolescents, the RIBS has presented acceptable psychometric performance. Chisholm et al., in 657 participants aged 11-13 years in a study, the RIBS showed Cronbach’s alpha of 0.86 (11). Mansfield et al., In a psychometric investigation with 1,032 adolescents between 11 and 15 years old, the instrument revealed a clear two-dimensional structure. The intended behaviors sub-scale showed high internal consistency, Cronbach’s alpha, and McDonald’s omega values of 0.94 (12).

Because the psychometric performance is specific for a given population, internal consistency and other measurements can vary over time and samples (13, 14). The performance of the Spanish version of RIBS is as yet unknown. However, given the need to quantify this problem in a developing country such as Colombia, its authors have attempted to establish frequencies for reported behavior and tested internal consistency and dimensionality for the intended behavior sub-scale in adolescent students (15).

For years, attribution theory has been used to explain the stigma-discrimination complex. Attribution theory deals with how behaviors are interpreted and the effect of those explanations on people’s perceptions (3). Besides, the theory holds that behavior is determined by a socially learned cognitive and emotional process (1). Thus, measuring mental health-related stigma-discrimination complex with a reliable and valid instrument has implications on anti-stigma education programs involving adolescents. It could make it possible to follow programs to reduce stigma, as a complex issue involving politics, economy, policies, and socio-historical processes.

The purpose of the study was to explore the performance of the RIBS among Colombian adolescents. This paper is the first study to evaluate the psychometric performance of the RIBS in a sample of participants in Colombia.

## SUBJECTS AND METHODS

A validation study was carried out. The Institutional Ethical Board reviewed and approved the project, parents signed informed consent, and adolescents agreed to participate. A non-probabilistic sample of 350 adolescent students was taken from two schools in Santa Marta, in northern Colombia. Santa Marta is a small Caribbean town of around 400,000 inhabitants. Participants were aged between 10 and 17 (*M*=13.2, *SD*=1.8). A total of 188 students (53.7%) were female, and 162 (46.3%) were male. Seventy students were in grade 6 (20.0%); 56 in 7 (16.0%); 59 in 8 (16.9%); 51 in 9 (14.6%); 70 in 10 (20.0%); and 44 in 11 (12.6%).

The participants completed the RIBS in the classroom. The instrument has two sub-scales with four items each. The first four items assess the prevalence of reported behaviors, whereas the latter four inquire about intended behaviors (10). The reported sub-scale offers two answer options: yes or no. The questions are listed in Table 1.

**Table 1.** Distribution of reported behaviors.

| Item | Yes<br>n (%) | No<br>n (%) |
| --- | --- | --- |
| Are you currently living with or have you ever lived with someone with a mental health problem? | 35 (10.0) | 315 (90.0) |
| Are you currently studying with or have you ever studied with someone with a mental health problem? | 61 (17.4) | 289 (82.6) |
| Do you currently have or have you ever had a neighbor with a mental health problem? | 87 (24.9) | 263 (75.1) |
| Do you currently have or have you ever had a close friend with a mental health problem? | 59 (16.9) | 291 (83.1) |

### Measurement

The intended sub-scale scores ranged from strongly agree (score 1) to strongly disagree (score 5). These questions are listed in Table 2. The scale followed a rigorous English to the Spanish translation process and a back translation by a professional translator to ensure semantic and cultural adaptation to Spanish spoken on the Colombian Caribbean coast.

**Table 2.** Distribution of participants’ responses for intended behaviors.

| Item | Strongly<br>agree<br>n (%) | Agree<br>n (%) | Neither<br>agree nor<br>disagree<br>n (%) | Disagree<br>n (%) | Strongly<br>disagree<br>n (%) |
| --- | --- | --- | --- | --- | --- |
| In the future, I would be willing to live with someone with a mental health problem | 31 (8.9) | 70 (20.0) | 146 (41.7) | 50 (14.3) | 53 (15.1) |
| In the future, I would be willing to work with someone with a mental health problem | 63 (18.0) | 125 (35.7) | 102 (29.1) | 32 (9.1) | 28 (8.0) |
| In the future, I would be willing to live nearby to someone with a mental health problem | 61 (17.4) | 129 (36.9) | 97 (27.7) | 31 (8.9) | 32 (9.1) |
| In the future, I would be willing to continue a relationship with a friend who developed a mental health problem | 69 (9.7) | 106 (30.3) | 102 (29.1) | 32 (9.1) | 41 (11.7) |

### Statistical analysis

Response frequencies and percentages were computed for the reported behavior sub-scale. Internal consistency reliability was quantified using Cronbach’s alpha and McDonald’s omega. McDonald’s omega deals with the bias resulting from the violation of the tau-equivalence principle when all items have a substantial and similar effect on reliability. Nevertheless, this assumption is constantly violated; thus, Cronbach’s alpha usually underestimates the internal consistency if tau-equivalence is not valid. Item-total score correlation and Cronbach’s alpha were calculated if the item was omitted. Finally, exploratory (EFA) and confirmatory factor analyses (CFA) were undertaken for the intended behavior sub-scale.

For EFA, the Kaiser-Meyer-Olkin measure of sampling adequacy (KMO), Bartlett’s Test of Sphericity (chi-squared), commonalities, loadings, and Eigenvalue (with explained variance) were computed. Moreover, for CFA, the calculated goodness-of-fit tests were chi-square test, with degrees of freedom (*df*) and probability value (*p*), RMSEA coefficient (Root mean square error of approximation of the approximation error) with 90% confidence interval (CI90%), Comparative Fit Index (CFI), Tucker-Lewis Index (TLI) and SMSR (Standardized mean square residual). These coefficients are acceptable when the chi-squared shows a probability value greater than 5%, RMSEA of below 0.06, CFI and TLI higher than 0.89, and SMSR lower than 0.05 (15). Analyses were carried out using Stata 13.0.

## RESULTS

The reported behaviors are presented in Table 1. Intended behavior sub-scale scores fell between 4 and 20 (*M*=10.8, *SD*=4.0). The pattern of answers for intended behaviors is presented in Table 2.

The Cronbach’s alpha was 0.88 (CI95% 0.86-0.90), and the McDonald omega, 0.88. KMO was 0.81; Bartlett chi-squared was 771.1 (*df*=6, *p*=0.01); and Eigenvalue 2.95 explained 73.9% of the total variance.

For the CFA, the Goodness-of-fit tests, chi squared was 21.9 (*df*=2, *p*=0.01); RMSEA, 0.17 (CI90% 0.11-0.24); CFI, 0.97; TLI, 0.92; and SMSR, 0.03. Other coefficients are presented in Table 3.

**Table 3.** Commonalities and loadings for intended behavior sub-scale.

| Item | Correlation *i-t* | Cronbach's alpha is the item is deleted | Commonality | Loading |
| --- | --- | --- | --- | --- |
| In the future, I would be willing to live with someone with a mental health problem | 0.73 | 0.85 | 0.58 | 0.76 |
| In the future, I would be willing to work with someone with a mental health problem | 0.77 | 0.83 | 0.76 | 0.87 |
| In the future, I would be willing to live nearby to someone with a mental health problem | 0.78 | 0.83 | 0.76 | 0.87 |
| In the future, I would be willing to continue a relationship with a friend who developed a mental health problem | 0.68 | 0.87 | 0.51 | 0.71 |

*i-t* corrected item-total score correlation.

## DISCUSSION

The present study illustrates the prevalence of mental disorder-related reported behaviors and psychometric performance of intended behaviors included in the RIBS. The prevalence of reported behaviors in this adolescent sample ranged from 10.0% to 24.9%. There is no information on these frequencies among adolescents, but the percentages of reported behaviors are lower than in adults. Evans-Lacko et al. found frequencies of between 24.8 and 43.9% in London, United Kingdom (10), Zalazar, et al., between 20.4 to 37.4% in Buenos Aires, Argentina (17). Similarly, college students report higher frequencies of reported behaviors than adolescent students. Yamaguchi et al. described a prevalence from 14.7 to 39.7% in Tokyo, Japan (18).

Likewise, this study documents high internal consistency for the intended behavior sub-scale. Cronbach’s alpha and McDonald’s omega were both 0.88. Previously, Pingani et al. and Yamaguchi et al. reported a Cronbach’s alpha of 0.83 (18, 19); Evans-Lacko et al. and Garcia et al. reported one of 0.85 (10, 20); Chisholm et al., of 0.86 (11); Aznar-Lou et al. reported a Cronbach’s alpha of 0.89 (21); and Mansfield et al., of 0.94 (12). Similarly, a high McDonald’s Omega has previously been reported, and Mansfield et al. found a coefficient of 0.94 (12). Clearly, the RIBS has shown high internal consistency in different populations.

Also, the one-dimensional intended behavior sub-scale was tested with a high Eigenvalue, which explains over 50% of the total variance. However, the CFA showed unsatisfactory values, lower than those usually expected for chi-squared and RMSEA. Evans-Lacko et al. omitted this information in their report (10). Similar to the present study’s findings, Garcia et al. reported poor indexes for chi-squared and RMSEA and acceptable values for CFI and TLI (20). Yamaguchi et al. found unsatisfactory values for chi-squared and RMSEA and high coefficients for CFI and TLI (18). However, Pingani et al. observed acceptable CFI and RMSEA, but they did not report chi-squared or TLI (19). These goodness-of-fit index findings are somewhat inconsistent.

The results corroborate the need for repeated reviews of the psychometric performance of construct measuring scales, such as the intended behavior sub-scale. Remarkable differences between populations frequently indicate limitations to construct validity and, as such, to all conclusions regarding the findings from a scale with a deficient performance (13, 14). The RIBS should be refined to guarantee its use in Spanish-speaking and adolescent populations (14, 22).

Besides, in this sample of Colombian adolescent students, the frequency of “strongly agree” for intended behaviors was relatively lower than for other adults; for instance, in the United Kingdom and Japan (10, 18). The findings corroborate the estimated high prevalence of mental disorder-related stigma-discrimination complex among the general Colombian population (23). Thus, asking for professional help in cases of mental disorder is much lower than the frequency of mental health problems, as they are defined -in the National Survey of Mental Health-, as symptoms that do not constitute a psychiatric problem (24).

Moreover, the mental disorder-related stigma-discrimination complex is a social determinant of mental health and a source of high costs in patients’ everyday lives; it reduces the use of services and discourages participation in leisure activities (9).

This study has two main strengths. It was the first time that a Spanish version of the RIBS was applied among adolescents. Furthermore, the second was that a broader analysis was carried out, including previously unreported CFA among adolescent students. Nevertheless, it is impossible to generalize these findings to other samples (13).

## CONCLUSIONS

To conclude, the RIBS can measure reported behaviors quickly, and the intended behavior sub-scale shows high internal consistency. However, the dimensionality of the intended behavior sub-scale presents modest goodness-of-fit indexes. These results need further replication in other Latin-American adolescent populations.

## Data Availability

All data produced in the present study are available upon reasonable request to the corresponding author.

## ACKNOWLEDGMENTS

The research was supported by the University of Magdalena, Santa Marta, and Human Behavioral Institute, Bogota, Colombia.

## Author contributions

A. Campo-Arias participated in the study, conceived the paper, conducted data analysis, wrote the manuscript, and approved the final version. G. A. Ceballos-Ospino participated in study design, oversaw data collection, analysis, interpretation, critically revised manuscript, and approved the final version. E. Herazo participated in study design, data analysis, data interpretation, and manuscript revision and approved the final version. The authors declare that they have no conflicts of interest.

## REFERENCES

1. Pescosolido BA, Martin JK. The stigma complex. Ann Rev Sociol 2015; 41: 87–116.

2. Arboleda-Flórez J. The rights of a powerless legion. In: Arboleda-Flórez J, Sartorius N. Understanding the stigma of mental illness: Theory and interventions. Chichester: John Wiley & Sons, Ltd.; 2008. P. 1–17.

3. Corrigan PW. Mental health stigma as social attribution: Implications for research methods and attitude change. Clin Psychol 2000; 7(1): 48–67.

4. Link BG, Phelan J. Stigma power. Soc Sci Med 2014: 103; 24–32.

5. Major B, O’Brien LT. The social psychology of stigma. Ann Rev Psychol 2005: 56: 393–421.

6. Campo-Arias A, Herazo E. [The stigma-discrimination complex associated with mental disorder as a risk factor for suicide]. Rev Colomb Psiquiatr. 2015; 44 (4): 243–50.

7. Phelan JC, Link BG, Dovidio JF. Stigma and prejudice: One animal or two? Soc Sci Med 2008; 67 (3): 358–67.

8. O’Driscoll C, Heary C, Hennessy E, McKeague L. Explicit and implicit stigma towards peers with mental health problems in childhood and adolescence. J Child Psychol Psychiatry. 2012; 53 (10): 1054–62.

9. Corrigan PW, Watson AC, Barr L. The self–stigma of mental illness: Implications for self– esteem and self–efficacy. J Soc Clin Psychol 2006; 25 (8): 875–84.

10. Evans-Lacko S, Rose D, Little K, Flach C, Rhydderch D, Henderson C, Thornicroft G. Development and psychometric properties of the reported and intended behaviour scale (RIBS): a stigma-related behaviour measure. Epidemiol Psychiatr Sci 2011; 20 (3): 263–71.

11. Chisholm K, Patterson P, Torgerson C, Turner E, Jenkinson D, Birchwood M. Impact of contact on adolescents’ mental health literacy and stigma: the SchoolSpace cluster randomised controlled trial. BMJ Open 2016; 6 (2): e009435.

12. Mansfield R, Humphrey N, Patalay P. Psychometric validation of the Reported and Intended Behavior Scale (RIBS) with adolescents. Stigma Health; 2020; 5 (3): 284–293.

13. Keszei AP, Novak M, Streiner DL. Introduction to health measurement scales. J Psychosom Res 2010; 68 (4): 319–33.

14. Ramada-Rodilla JM, Serra-Pujadas C, Delclós-Clanchet GL. Cross-cultural adaptation and health questionnaires validation: revision and methodological recommendations. Salud Publica Mex 2013: 55 (1): 57–66.

15. Hu LT, Bentler PM. Cutoff criteria for fit indexes in covariance structure analysis: Conventional criteria versus new alternatives. Struct Equat Model 1999; 6 (1):1–55.

16. Khandelwal S, Avodé G, Baingana F, Conde B, Cruz M, Deva P, et al. Mental and neurological health research priorities setting in developing countries. Soc Psychiatry Psychiatr Epidemiol 2010: 45 (4):487–95.

17. Zalazar V, Leiderman EA, Agrest M, Nemirovsky M, Lipovetzky G, Thornicroft G. Reported and intended behavior towards people with mental health problems in Argentina. Int J Ment Health 2018; 47 (3): 215–27.

18. Yamaguchi S, Koike S, Watanabe KI, Ando S. Development of a Japanese version of the Reported and Intended Behaviour Scale: Reliability and validity. Psychiatry Clin Neurosci 2014; 68 (6): 448–55.

19. Pingani L, Evans-Lacko S, Luciano M, Del Vecchio V, Ferrari S, Sampogna G, et al. Psychometric validation of the Italian version of the Reported and Intended Behaviour Scale (RIBS). Epidemiol Psychiatr Sci 2016; 25 (5): 485–92.

20. Garcia C, Golay P, Favrod J, Bonsack C. French translation and validation of three scales evaluating stigma in mental health. Front Psychiatry 2017; 8: 290.

21. Aznar-Lou I, Serrano-Blanco A, Fernández A, Luciano JV, Rubio-Valera M. Attitudes and intended behaviour to mental disorders and associated factors in Catalan population, Spain: cross-sectional population-based survey. BMC Public Health 2016; 16: 127.

22. Schmitt TA. Current methodological considerations in exploratory and confirmatory factor analysis. J Psychoeduc Assess 2011; 29 (4): 304–21.

23. Ceballos GA, Jiménez MP., De la Torre H., Colorado YS. [Stigma and discrimination in medical students towards people with mental disorders]. Tesis Psicol 2020; 15 (2): 238–251.

24. Ministerio de Salud de Colombia-Colciencias. National Mental Health Survey 2015. Bogota: Javegraf; 2015.

